# Accuracy of Expired BinaxNOW Rapid Antigen Tests

**DOI:** 10.1101/2023.05.17.23290131

**Authors:** Mary Jane E. Vaeth, Omar Abdullah, Minahil Cheema, Kristie Sun, Maryam Elhabashy, Asia Mitchell, Maisha Foyez, Mahita Talla, Aamna Cheema, Charles Locke, Melinda Kantsiper, Andrew Pekosz, Heba H. Mostafa, Zishan K. Siddiqui

## Abstract

The widespread existence of expired antigen testing kits in households and potential coronavirus outbreaks necessitate evaluating the reliability of these expired kits. Our study examined BinaxNOW COVID-19 rapid antigen tests 27 months post-manufacture and 5 months past their FDA extended expiration dates, using SARS-CoV-2 variant XBB.1.5 viral stock. We conducted testing at two concentrations: the Limit of Detection (LoD) and 10 times the LoD. 100 expired and unexpired kits were tested at each concentration for a total of 400 antigen tests. At the LoD (2.32x10^2 TCID50/mL), both expired and unexpired tests displayed 100% sensitivity (95% CI 96.38% to 100%), with no statistical difference (95% CI -3.92% to 3.92%). Similarly, at 10 times the LoD, unexpired tests retained 100% sensitivity (95% CI 96.38% to 100%), while expired tests exhibited 99% sensitivity (95% CI 94.61% to 99.99%), demonstrating a statistically insignificant 1% difference (95% CI -2.49% to 4.49%, p=0.56). Expired rapid antigen tests had fainter lines than the unexpired tests at each viral concentration. The expired rapid antigens tests at LoD were only just visible. These findings carry significant implications for waste management, cost efficiency, and supply chain resilience in pandemic readiness efforts. They also provide critical insights for formulating clinical guidelines for interpreting results from expired kits. In light of expert warnings of a potential outbreak of a severity rivaling the Omicron variant, our study underscores the importance of maximizing the utility of expired antigen testing kits in managing future health emergencies.

## Background

In the wake of the COVID-19 pandemic, rapid antigen tests have emerged as a vital tool in the global response to contain virus spread^1^. In the rush to make these tests available, the authorized shelf life had been set at only about four to six months from the day of manufacture.^2^ Despite the short expiration dates, these tests have provided a fast, accessible, and cost-effective alternative to the gold standard polymerase chain reaction (PCR) testing.^3^ Rapid antigen tests detect viral proteins and offer the advantages of shorter turnaround times, increased testing capacity, and broad-scale screening for mitigating the risk of transmission.^4^ While PCR tests remain superior in terms of sensitivity, rapid antigen tests have proven invaluable in identifying infectious individuals^5,6,7^ particularly during the acute phase of infection, thus playing a crucial role in curbing the pandemic.

Recognizing the importance of accurate expiration date determination is crucial for preventing wastage of precious resources, maintaining preparedness in the face of emerging variants, and strengthening the resilience of the supply chain for diagnostic tests.^8^ As global demand for rapid antigen tests continues, maximizing their utility and minimizing waste are critical in ensuring that healthcare systems worldwide can continue to rely on these tests for swift and efficient public health interventions.^9^ The expiration dates for COVID-19 rapid antigen tests were initially determined based on accelerated stability studies, which are commonly used to estimate the shelf life of diagnostic products.^10^ In light of real-world data and ongoing stability testing, the FDA and other regulatory bodies have extended the expiration dates of some rapid antigen tests, allowing for their continued use beyond the initial estimates.^11^

Updates to the expiration dates of rapid antigen kits have necessitated a broader public health campaign to alert the general public to the potential inaccuracy of these dates. Recommendations regarding these kits vary across public health agencies and health centers. The FDA, for instance, advises checking the expiration date on its webpages and using the kits within the extended expiration date provided by the FDA. Some entities, recommend discarding the kits if they are listed as expired on the FDA webpage.^12^ In contrast, the California Department of Public Health supports the emergency use of over-the-counter COVID-19 tests beyond their FDA-authorized expiration, provided that the internal control line remains both easily visible and of the color specified by the test instructions after test development.^13^

The high prevalence of unused and expired antigen testing kits in households, often used past their expiration dates due to convenience or unawareness, necessitates clearer clinical guidance on interpreting results from such kits.^14^ Leveraging these kits to their maximum usable lifespan could enhance waste management, cost efficiency, and supply chain robustness in pandemic readiness efforts. This urgency is amplified by recent expert warnings received by the White House, indicating a roughly 20% chance of a coronavirus outbreak in the next two years, potentially rivaling the severity of the Omicron variant.^15^ There is limited peer-reviewed information available about the accuracy of expired kits. It has been shown that they work well when robust antigen laden “positive controls” are used to assess their accuracy,^16^ but more thorough analysis data is not available. In response to this knowledge gap, our study aims to evaluate the performance of expired kits at the lower bound of their detection capability, to inform future public health decision-making.

## Methods

Our study investigated the accuracy of expired BinaxNOW rapid antigen tests at or above the Limit of Detection (LoD) as compared with unexpired tests. We began by determining the LoD for the unexpired test using our prior study as a reference point (ref). The unexpired BinaxNOW tests used in the study had the following characteristics: lot #182848, manufacturer date 18 February 2022, and FDA extended expiration date 1 October 2023. The expired BinaxNOW kits used in the study had lot #134545, manufacturer date 25 November 2020, and FDA extended expiration date 16 September 2022.^17^ The tests kits were studied in March 2023.

SARS-CoV-2 variant XBB.1.5 viral stock (HP40900, EPI_ISL_16026423) isolated from a clinical sample and quantified at 2.32x10^6 TCID50/mL^18^ was used to make serial dilutions in Universal Viral Transport Media (UTM/VTM). Rapid antigen testing was performed by dipping the swab in the UTM/VTM dilutions, followed by testing per manufacturer’s instructions. The LoD was determined using unexpired swabs and a 1 log higher (i.e. 10x times higher) concentration was used to test both expired and unexpired swabs side by side. For each set of 100 expired and 100 unexpired tests, we conducted rapid antigen testing at the concentration 10 times above the LoD. Additionally, we conducted rapid antigen testing for another set of 100 expired and 100 unexpired tests at the LoD concentration as secondary analysis. In total, we tested 400 rapid antigen tests. We used the manufacturer-provided swabs to obtain samples from the diluted viral concentrations and followed the manufacturer’s instructions for testing. The sensitivity of the expired and unexpired rapid antigen tests was calculated at each concentration. Chi-square test was used to sample proportions where possible.

## Results

The expired kits were tested 27 months after their manufacture and 5 months beyond their FDA extended expiration dates. The unexpired kits were tested 13 months after manufacture and had over 6 months remaining until expiration after most recent FDA extensions. At concentrations 10xLoD (2.32x10^3 TCID50/mL), 100/100 (Sensitivity 100%, 95% CI 96.38% to 100%) of the unexpired rapid antigen tests tested positive and 99/100 (Sensitivity 99%, 95% CI 94.61% to 99.99%) of the expired tests tested positive. This represented a statistically insignificant difference of 1% in sensitivity compared with unexpired tests (95% CI -2.49% to 4.49%, p=0.56). Similarly, at a concentration of LoD (2.32x10^2 TCID50/mL), both the expired and unexpired tests detected 100/100 (Sensitivity 100%, 95% CI 96.38% to 100%) virus samples. The difference between the unexpired and expired tests was 0% (95% CI -3.92% to 3.92%) (Table 1). Seven unexpired rapid antigen kits were tested at 2.32x10 TCID50/mL concentration and all were negative.

**Table 1:**
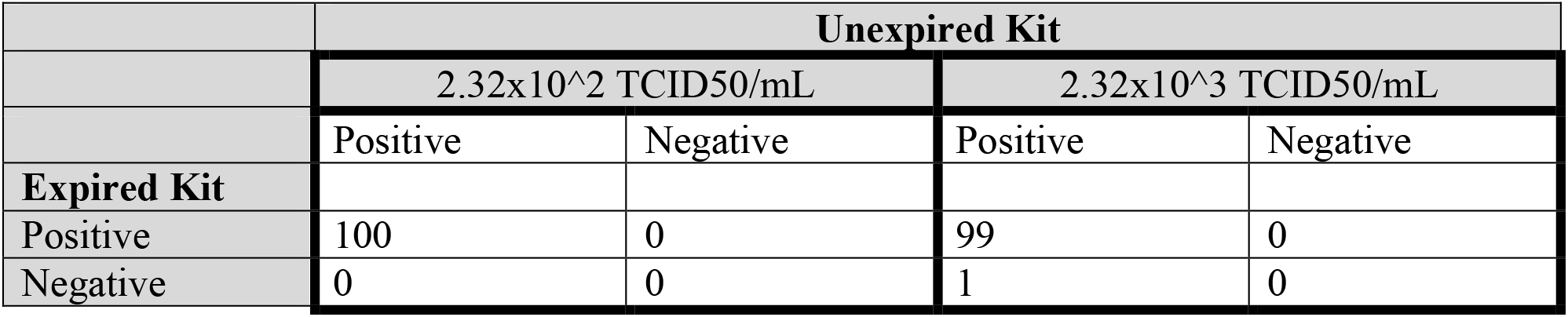
Agreement Matrix for Expired and Unexpired Rapid Antigen Test using viral concentration at LoD and at 10xLoD

|  | Unexpired Kit |  |  |  |
| --- | --- | --- | --- | --- |
|  | 2.32x10<sup>2</sup> TCID<sub>50</sub>/mL |  | 2.32x10<sup>3</sup> TCID<sub>50</sub>/mL |  |
|  | Positive | Negative | Positive | Negative |
| Expired Kit |  |  |  |  |
| Positive | 100 | 0 | 99 | 0 |
| Negative | 0 | 0 | 1 | 0 |

Qualitatively, the sample and control lines were fainter with the expired kits than with the unexpired kits and for the lower concentration compared to the higher concentration. The sample line with low concentration virus sample for the expired kits was barely visible, but this was consistent through all 100 expired tests at this concentration (Figure 1).

**Figure 1:**
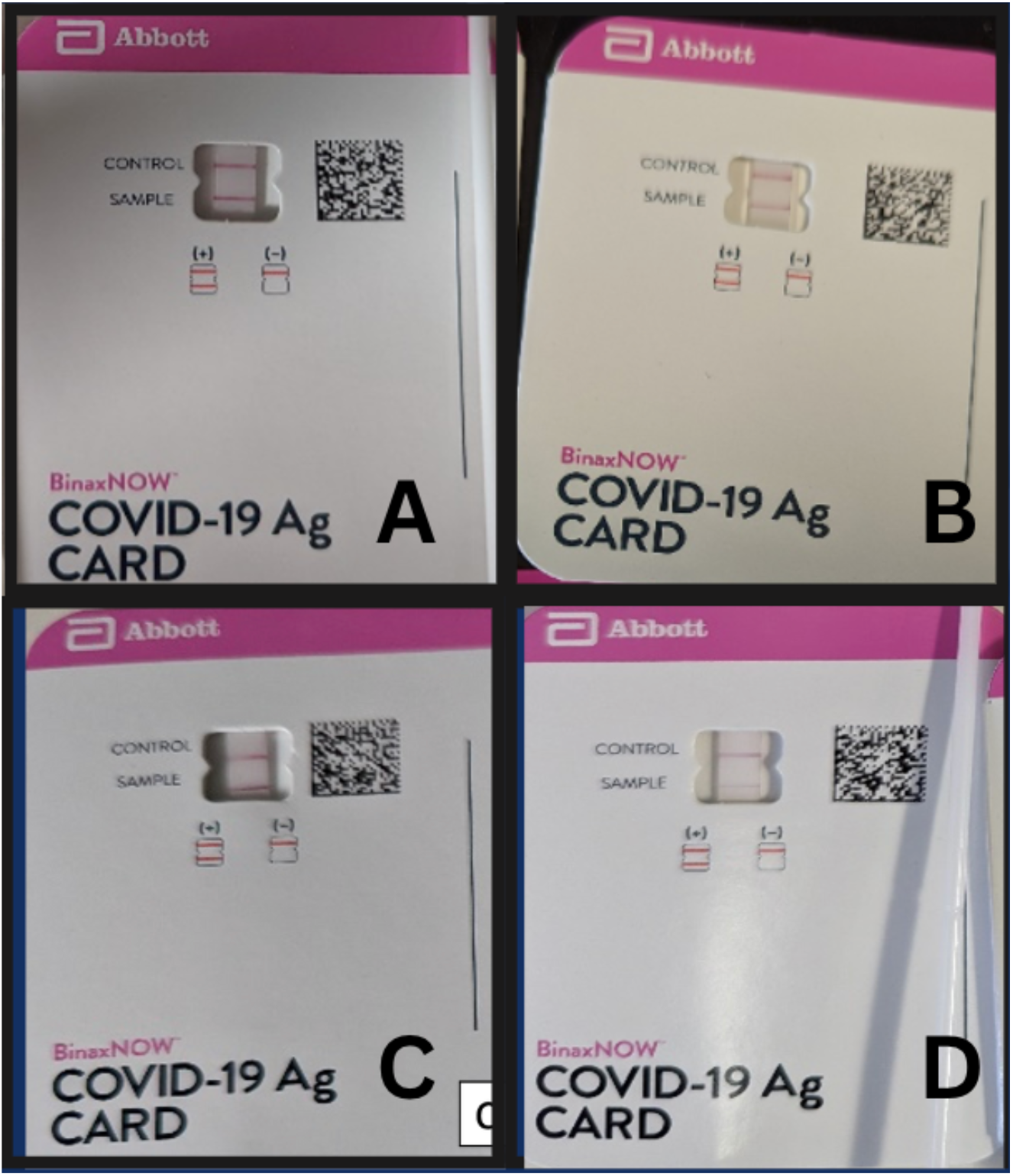
Unexpired and Expired Rapid Antigen Kits at varying viral concentrations **A. Unexpired 2.32X10^3 TCID50/mL (10xLoD)** **B. Unexpired 2.32X10^2 TCID50/mL (LoD)** **C. Expired 2.32X10^3 TCID50/mL (10xLoD)** **D. Expired 2.32X10^2 TCID50/mL (LoD)**

## Discussion

Our study aimed to evaluate the performance of expired COVID-19 rapid antigen tests, specifically the BinaxNOW kits, in comparison to their unexpired counterparts at the lower bound of their detection capability. The findings suggest that there is no statistically significant difference in the sensitivity of expired and unexpired rapid antigen tests at both the concentration 10 times above the LoD and at the LoD with SARS-CoV-2 variant XBB.1.5. This is an important observation as it indicates that expired rapid antigen tests may still be a useful tool in detecting SARS-CoV-2 infections, particularly during periods of high demand or supply chain disruptions.

The results of our study are in line with recent extensions of expiration dates by the FDA and other regulatory bodies, which have been based on real-world data and ongoing stability testing. These extensions have helped to reduce waste and improve the overall efficiency of the diagnostic testing supply chain. Our findings support this. Our study goes a step further in supporting that some expired tests may still have utility beyond their extended expiration dates. However, it is essential to note that the qualitative assessment of the sample and control lines revealed that they were fainter with the expired kits compared to the unexpired kits. This observation may have implications on the user’s ability to accurately interpret the results, particularly in low-resource settings or for individuals with limited experience in performing and interpreting rapid antigen tests. This aspect warrants further investigation and highlights the need for clear guidance on interpreting test results from expired kits.

While our study provides valuable insights into the performance of expired rapid antigen tests, it has some limitations. First, our study focused solely on the BinaxNOW rapid antigen test, and the findings may not be generalizable to other rapid antigen tests available in the market. Second, the study’s sample size is relatively small, and future studies with larger sample sizes for tests stored in different conditions may provide more robust estimates of the sensitivity of expired tests. Lastly, the study was conducted under controlled laboratory conditions, which may not fully represent real-world conditions. Factors like humidity, temperature, storage conditions, and user variability can also impact the test performance. However, despite these limitations, our study provides essential information on the potential utility of expired rapid antigen test kits at the lower bound of detection, which can be considered in public health decision-making processes.

In conclusion, our study suggests that some expired BinaxNOW COVID-19 rapid antigen tests may still offer similar sensitivity to currently circulating SARS-CoV-2 variants when compared to their unexpired counterparts, though with potential limitations in the visibility of the test lines. These findings can inform public health decision-making, particularly in situations where resources are limited, and access to testing is crucial for containing the spread of the virus. Future research should explore the performance of other rapid antigen test brands beyond their expiration dates and investigate accuracy in real-world settings. Our findings have significant implications for public health policy, as they support the potential for more flexible expiration date guidelines, which could minimize test kit wastage and strengthen diagnostic test supply chains during the ongoing COVID-19 pandemic and future public health crises.

## Data Availability

All data produced in the present work are contained in the manuscript.

